# Validation of an AI-powered mobile application for personalizing medical note explanations

**DOI:** 10.1101/2025.09.17.25335707

**Authors:** Nicholas Lamb

## Abstract

Almost half of adults struggle to understand written health information, making medical communication a critical barrier to patient care. While AI shows promise for improving health communication, few tools have been rigorously validated for personalizing medical explanations. I developed Patiently AI, a mobile application that uses large language models to simplify medical notes with audience-specific adaptations (child, teenager, adult, carer) and tone variations (friendly, informative, reassuring). Our three-phase validation comprised: computational analysis of readability improvements across 210 AI-generated outputs using established metrics; expert evaluation by 15 healthcare professionals assessing clinical accuracy, safety, and communication quality; and patient survey of 54 participants evaluating preferences, comprehension, and acceptance. AI-generated explanations showed significant readability improvements: mean Flesch–Kincaid Grade Level decreased by 2.96 levels (10.57→7.61), Flesch Reading Ease increased by 31.9 points (37.7→69.6), and Gunning Fog Index decreased by 4.09 points (14.5→10.4). Improvements were greatest for younger audiences (child: 4.25 grade level reduction vs. adult: 1.80). Expert evaluation rated AI outputs highly for medical accuracy (4.49±0.51/5), clarity (4.53±0.50/5), and trustworthiness (4.37±0.58/5), with 87.3% deemed clinically safe. Patient evaluation showed strong acceptance: 70.0% preferred AI-generated explanations, with high ratings for clarity (4.58±0.52/5) and confidence in care (4.19±0.65/5). 70.4% of patients indicated likelihood to use the application. This study provides robust evidence that AI can safely and effectively personalize medical communication while maintaining clinical accuracy. The validated Patiently AI application represents a scalable solution for improving health literacy and patient engagement across diverse populations.

## Introduction

Health literacy – the capacity to obtain, process, and understand basic health information needed to make appropriate health decisions^1^ – remains a persistent challenge in healthcare delivery. Almost half of adults struggle to understand written health information^2^, with disproportionate impacts on older adults, ethnic minorities, and those with lower educational attainment^3^. Poor health literacy is associated with increased hospitalizations, emergency department visits, medication errors, and mortality^4,5^, with estimated annual costs exceeding $236 billion.^6^

The 21st Century Cures Act’s mandate for immediate patient access to medical records has amplified this challenge, as patients now receive complex documentation without interpretation or simplification^7^. Medical documentation, while essential for clinical care, often presents significant comprehension barriers for patients. Clinical notes contain complex medical terminology, abbreviations, and technical language that can impede patient understanding and engagement^8^. Traditional approaches to improving medical communication, such as plain language guidelines and health literacy training, have shown modest benefits but face scalability challenges^9,10^.

The emergence of large language models (LLMs) presents unprecedented opportunities to personalize health communication at scale^11,12^. Recent studies have demonstrated AI’s potential to simplify medical text^13,14^, translate complex information^15^, and adapt communication styles^16^. However, most existing research focuses on general text simplification rather than medical-specific applications, and few studies have rigorously evaluated AI-generated health communications for clinical accuracy, safety, and patient acceptance^17^.

Recent evidence highlights both the promise and perils of AI-generated patient communications. While Stanceski et al. (2024) demonstrated that AI-generated discharge instructions achieved better readability and language simplification compared to original summaries, safety issues were identified in 18% of cases, including 6% with hallucinations and 3% containing new medications not present in the original discharge summaries^18^. Furthermore, effective health communication requires more than simple text reduction – it demands audience-appropriate adaptation that considers age, health literacy level, and relationship to care (patient vs. caregiver)^19,20^. Children, teenagers, adults, and caregivers have distinct communication needs, preferred language styles, and information processing capabilities^21,22^. The emotional context of medical information – whether informative, reassuring, or friendly – significantly impacts patient engagement and understanding^23^. Current AI applications largely employ one-size-fits-all approaches that fail to address this diversity.

To address these gaps, I developed Patiently AI, a mobile application that uses advanced language models to transform medical notes into personalized, audience-appropriate explanations. The application adapts both content complexity and communication tone based on user-specified parameters, creating tailored explanations for different audiences (child, teenager, adult, carer) and communication contexts (friendly, informative, reassuring).

This study presents a comprehensive validation of Patiently AI, a commercially deployed iOS application, through three complementary evaluation approaches: computational readability analysis, expert clinical assessment, and patient preference evaluation. Our objectives were to: (1) quantify readability improvements across different audience adaptations; (2) assess clinical accuracy, safety, and communication quality through expert review; and (3) evaluate patient preferences, comprehension, and acceptance of AI-generated explanations.

## Methods

### Study design

I carried out a prospective, mixed-methods evaluation study with three distinct components designed to comprehensively assess the Patiently AI application across technical, clinical, and user dimensions. The study was registered with the Open Science Framework (OSF.io/3CDX6) and conducted in accordance with CONSORT-AI guidelines for reporting AI interventions^24^.

### Intervention

Patiently AI is powered by advanced large language models (GPT-4o, OpenAI) with specialized prompt architectures designed for healthcare communication. The app accepts text or voice input and generates simplified versions based on audience (Adult/Teenager/Child/Caregiver) and tone (Informative/Friendly/Reassuring) parameters, supporting 11 languages without storing personal health information. Each combination employs specific linguistic strategies, vocabulary levels, and explanation approaches optimized for the target audience.

### Study 1: Technical validation

We generated 10 synthetic medical notes using AI to simulate common clinical scenarios covering hypertension, diabetes, mental health, oncology, cardiology, neurology, and radiology. These notes were designed to reflect typical medical documentation complexity and terminology. Each medical note was processed through Patiently AI to generate 7 audience-tone combinations. Each combination was processed in triplicate to account for variability in AI-generated outputs, totaling 210 simplifications (10 notes × 7 combinations × 3 replicates). We compared these against the original medical notes using three established readability metrics:

- Flesch-Kincaid Grade Level (FKGL): Estimates the U.S. school grade level required for comprehension
- Flesch Reading Ease: Scores text from 0 (very difficult) to 100 (very easy) based on sentence length and syllable count
- Gunning Fog Index: Estimates years of formal education needed to understand text on first reading

Readability calculations were assessed using the online Readability Formulas tool (https://readabilityformulas.com).

### Study 2: Expert clinical review

Healthcare professionals were recruited through professional networks and the Prolific research platform. Each received 10 AI-simplified outputs via an online survey, rating clinical safety (binary), preservation of medical facts, clarity, completeness, tone appropriateness, and trustworthiness (5-point Likert scales). Experts also provided qualitative feedback on strengths, concerns, and suggestions for improvement.

Mean scores and standard deviations were calculated for each evaluation criterion. Safety assessments were analyzed as proportions with 95% confidence intervals. Qualitative feedback was analyzed thematically to identify common patterns and concerns.

### Phase 3: Patient evaluation

We recruited participants through the Prolific research platform, which provides access to diverse, pre-screened research participants. Inclusion criteria required English fluency and age ≥18 years. Participants evaluated medical note pairs (original vs. AI-simplified) across 5 clinical scenarios, indicating preference, completing comprehension questions, and rating clarity and confidence. The survey concluded with questions about likelihood to use Patiently AI and general feedback.

Mean preference scores, satisfaction ratings, and usage intentions with 95% confidence intervals were calculated. Preference scores ≥4 were classified as “preferring AI-generated explanations.” Descriptive statistics characterized participant demographics and response patterns.

## Results

### Participant characteristics

The study included 54 patients (98% completion rate) and 15 expert reviewers. The patient sample was 51.9% male, predominantly middle-aged (40.7% aged 30–49 years), 66.7% with university education, 74.1% reporting chronic health conditions, and 25.9% identifying as a caregiver. The expert panel comprised: medical writers (n=4), general practitioners (n=2), specialists (n=2), clinical researchers (n=2), pharmacists (n=1), physiotherapists (n=1), and other healthcare professionals (n=3). Mean experience was 8.3 years (range: 1–15+ years).

### Readability improvements

AI-generated explanations demonstrated substantial readability improvements across all metrics (Table 1). Mean Flesch–Kincaid Grade Level decreased from 10.57 to 7.61 (improvement: 2.96 grade levels, 95% CI: 2.64–3.28). Flesch Reading Ease increased from 37.7 to 69.6 (improvement: 31.9 points, 95% CI: 29.8–34.0). Gunning Fog Index decreased from 14.5 to 10.4 (improvement: 4.09 points, 95% CI: 3.76–4.42).

**Table 1.** Readability metrics comparison

| Metric | Original notes mean (SD) | AI-generated mean (SD) | Improvement mean (95% CI) | P value |
| --- | --- | --- | --- | --- |
| Flesch–Kincaid Grade Level | 10.57 (2.84) | 7.61 (1.98) | 2.96 (2.64–3.28) | <0.001 |
| Flesch Reading Ease | 37.69 (15.42) | 69.60 (12.88) | 31.90 (29.8–34.0) | <0.001 |
| Gunning Fog Index | 14.49 (3.67) | 10.40 (2.45) | 4.09 (3.76–4.42) | <0.001 |

Readability improvements varied significantly by target audience (Fig. 1). Child-focused explanations showed the greatest improvements (FKGL reduction: 4.25 grade levels), followed by teenager (3.58 grade levels), adult (1.80 grade levels), and carer-focused explanations (1.64 grade levels). This gradient reflects the application’s ability to systematically adapt complexity based on intended audience developmental and literacy characteristics.

**Fig. 1.**
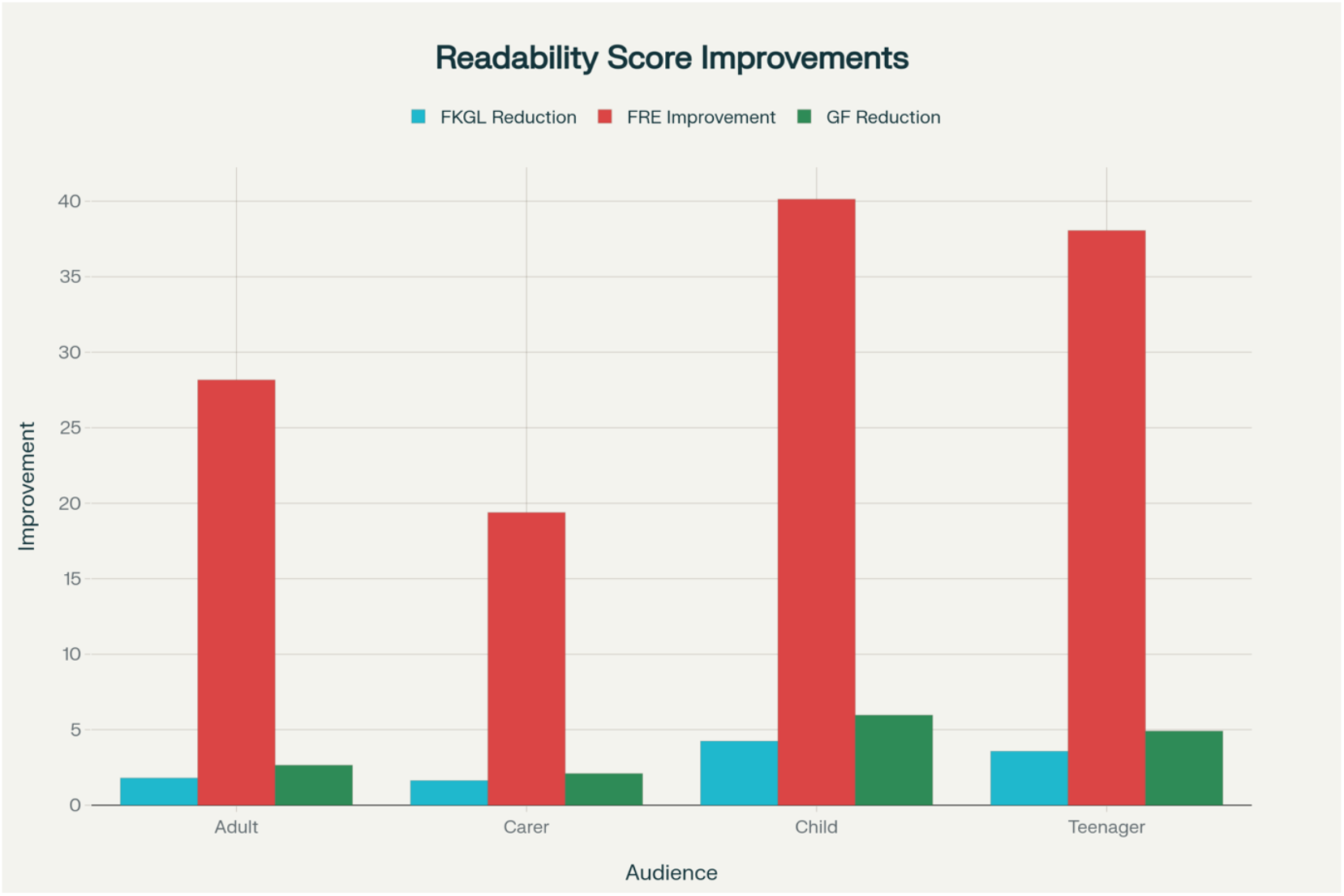
Readability improvements by target audience. Child explanations resulted in the largest reduction in Flesch-Kincaid Grade Level (FKGL; 4.25), greatest improvement in Flesch Reading Ease (FRE; 40.1 points), and the largest drop in Gunning Fog Index (GF; 6.0 points) compared to Adult, Teenager, and Carer adaptations.

Original medical notes averaged a college-level reading requirement (FKGL: 10.57), placing them beyond the comprehension level of many patients. AI-generated explanations achieved middle school reading levels on average (FKGL: 7.61), with child-focused explanations reaching elementary school levels (FKGL: 6.32).

### Expert Clinical Evaluation

Healthcare professionals (n=15) provided highly favorable evaluations of AI-generated explanations across all assessment dimensions (Table 2). Medical accuracy received the highest rating (4.49±0.51/5), followed by clarity (4.53±0.50/5), completeness (4.39±0.69/5), tone appropriateness (4.37±0.72/5), and trustworthiness (4.37±0.58/5).

**Table 2.** Expert clinical review ratings

| Assessment criterion | Mean score (SD) | Range | High rating (≥4) |
| --- | --- | --- | --- |
| Medical accuracy | 4.49 (0.51) | 3.0–5.0 | 93.3% |
| Clarity | 4.53 (0.50) | 3.0–5.0 | 94.7% |
| Completeness | 4.39 (0.69) | 2.0–5.0 | 86.7% |
| Tone appropriateness | 4.37 (0.72) | 2.0–5.0 | 84.0% |
| Trustworthiness | 4.37 (0.58) | 2.0–5.0 | 86.0% |
| Clinical Safety | - | - | 87.3% safe |

Clinical safety assessment revealed that 87.3% of AI-generated explanations (131/150) were deemed clinically safe, with 12.7% (19/150) flagged for potential safety concerns. Common safety concerns included: overly simplified explanations that might minimize serious conditions (n=8), addition of general advice not present in original notes (n=6), and potential for patient misinterpretation (n=5).

Qualitative feedback from experts highlighted several strengths. One expert described the outputs as “*well written, clear and understandable by most people*”. Others highlighted the “*good level of detail and reassurance*” and noted the explanations “*preserve the key medical facts*” while “*meeting the intended tone and audience*”. However, some experts noted, “*Although it covers everything, it is too much to read*… *Assurance does need words, but if it can be less lengthy to read*.”

### Patient preference and acceptance

Patient evaluation (n=54) demonstrated strong preference for AI-generated explanations (Table 3). Overall preference scores averaged 3.89/5 across scenarios, with 70.0% of responses indicating preference for AI-generated explanations (score ≥4). Preference varied by clinical scenario, ranging from 3.41/5 (mental health, child audience) to 4.30/5 (post-COVID symptoms, adult audience).

**Table 3.** Patient Preference and Satisfaction Results

| Outcome Measure | Mean score (SD) | Range | Positive Response (≥4) |
| --- | --- | --- | --- |
| Overall Preference | 3.89 (1.22) | 1.0–5.0 | 70.0% |
| Clarity Rating | 4.58 (0.52) | 3.0–5.0 | 96.3% |
| Confidence in Care | 4.19 (0.65) | 3.0–5.0 | 78.5% |
| Likelihood to Use | 3.83 (1.35) | 1.0–5.0 | 70.4% |

All ratings use 1–5 Likert scales where 5 is the most positive response.

Clarity ratings were consistently high across all scenarios (mean: 4.58±0.52/5), indicating that participants found AI-generated explanations easy to understand. Confidence in care ratings also showed positive results (mean: 4.19±0.65/5), suggesting that simplified explanations enhanced rather than diminished patient confidence in their medical care.

Comprehension assessment through scenario-specific questions revealed high accuracy rates (mean: 94.3% correct responses), confirming that readability improvements did not compromise information transfer. Participants successfully identified key information including medications, follow-up timing, and care instructions.

Usage intentions were favorable, with 70.4% of participants indicating likelihood to use Patiently AI for simplifying their medical notes (score ≥4/5). Mean usage likelihood was 3.83/5, suggesting broad acceptance of AI-powered medical communication tools.

### Audience-specific performance

Analysis by target audience revealed systematic differences in both readability improvements and user acceptance. Child-focused explanations achieved the greatest readability gains but received mixed expert feedback regarding age-appropriateness for serious medical conditions. Teenager-focused explanations balanced readability with clinical seriousness effectively. Adult-focused explanations maintained professional tone while improving accessibility. Carer-focused explanations emphasized actionable guidance and support resources.

Patient preferences showed scenario-dependent variations, with adult-focused explanations receiving consistently high ratings across diverse clinical contexts. Child and teenager explanations were well-received for appropriate clinical scenarios but less preferred for serious conditions requiring more careful communication. These patterns are illustrated in Fig. 2, which shows that over 70% of patients preferred the AI explanations in three out of five scenarios, with mean ratings above 4.0 (on a 1–5 scale) in three scenarios. The highest preference was for scenario 5 (mean 4.30, 85.2% preferring AI explanation).

**Fig. 2.**
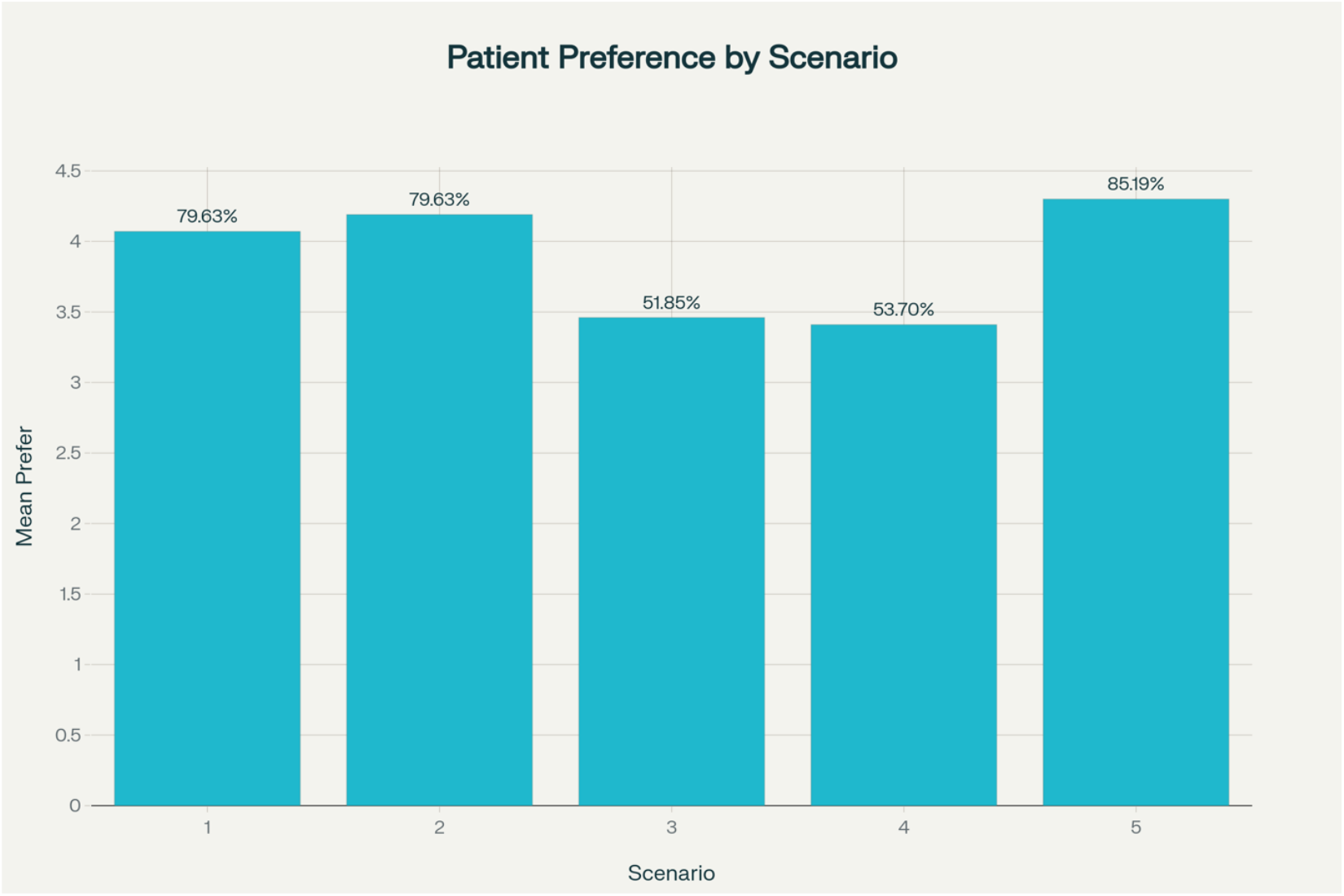
Patient preference for AI explanations. Patient ratings of AI-generated explanations were measured for five real-world clinical scenarios: (1) hypertension management (ramipril added for persistently high blood pressure); (2) diabetes management (consideration of GLP-1 therapy for persistently elevated HbA1c); (3) child mental health review (sertraline initiated for mood and sleep, no suicidal ideation); (4) chronic migraine management (topiramate and headache diary); and (5) referral for ongoing post-COVID breathlessness.

## Discussion

This comprehensive validation study provides robust evidence that AI can safely and effectively personalize medical communication while maintaining clinical accuracy. The Patiently AI application demonstrated significant readability improvements, high expert ratings for clinical quality, and strong patient acceptance across diverse medical scenarios.

The observed readability improvements are clinically meaningful. Reducing reading level requirements by nearly 3 grade levels transforms medical notes from college-level text to middle school accessibility, potentially benefiting millions of patients with limited health literacy. The systematic variation by audience type demonstrates sophisticated AI adaptation capabilities that go beyond simple text reduction to include audience-appropriate language, concepts, and communication styles. These improvements align with health literacy recommendations that suggest optimal health materials should target 6th-8th grade reading levels^25^. Our results show AI-generated explanations achieved this target while preserving essential medical information, addressing a key challenge in health communication.

Expert evaluation revealed high confidence in AI-generated explanations’ clinical safety and accuracy. In this study, 87.3% of AI-generated explanations were rated safe by the expert panel. While encouraging, this highlights the need for continued vigilance in AI medical applications. The 12.7% of explanations flagged for safety concerns primarily involved over-simplification of serious conditions rather than factual errors, suggesting areas for prompt engineering refinement rather than fundamental technical limitations.

The strong expert ratings for medical accuracy (4.49/5) and completeness (4.39/5) indicate that AI successfully preserved critical medical information while improving accessibility. This addresses concerns that simplification necessarily compromises clinical accuracy – a key barrier to adoption of health literacy interventions.

Our findings align with recent implementation evidence from Shah et al. (2025), who demonstrated that clinicians using AI-generated draft explanations for laboratory, imaging, and pathology results reported improved efficiency (71.0%) and higher quality patient explanations (72.0%). However, their study also identified challenges with content accuracy and completeness, emphasizing the need for continued refinement of AI-generated medical communications.^26^

High patient preference scores (mean 70% preferring AI explanations across scenarios) and strong clarity ratings (4.58/5) demonstrate genuine user value beyond technical metrics. Importantly, improved comprehension corresponded with increased confidence in care (4.19/5), suggesting that better understanding enhances rather than undermines trust in medical recommendations. The 70.4% expressing likelihood to use the application indicates market readiness and potential for real-world adoption. This acceptance rate compares favorably with other digital health tools and suggests broad patient interest in AI-powered health communication support^27^.

These findings have several important implications for healthcare delivery. First, unlike traditional health literacy interventions requiring extensive provider training or resource allocation, AI-powered tools can provide personalized communication support at scale. Second, rather than replacing provider-patient communication, AI explanations can prepare patients for more effective clinical encounters by improving baseline understanding. Third, audience-appropriate adaptations may help address health communication disparities affecting vulnerable populations. Fourth, the mixed expert feedback suggests that AI medical communication tools benefit from clinical oversight, particularly for serious diagnoses or vulnerable populations.

Several study limitations warrant consideration. First, the expert panel, while diverse in background, was relatively small (n=15) and may not represent all clinical perspectives. Larger, more diverse expert evaluations would strengthen confidence in safety assessments. Second, patient evaluation occurred in a controlled survey environment rather than real clinical contexts. Actual usage patterns, comprehension outcomes, and care impacts may differ from survey responses. Longitudinal studies tracking real-world usage and health outcomes are needed. Third, this study used AI-generated medical notes for validation, which ensured patient privacy and allowed systematic testing across diverse clinical scenarios. However, future validation with authentic clinical documentation will strengthen generalizability to real-world contexts, where formatting and complexity may differ. Finally, our evaluation focused on English-language content with predominantly educated participants. Generalizability to other languages, cultural contexts, or lower health literacy populations requires additional validation.

The ethical implications of our work must also be considered within the broader context of patient AI use. Pal et al.’s (2025) comprehensive review of 44 studies found that while AI-generated medical information is generally accurate and readable, significant risks remain including lack of transparency regarding information sources and potential for ‘humanizing’ AI models that may lead to overreliance. Their findings reinforce the importance of appropriate governance and clinician oversight in AI-mediated patient communication tools^28^.

The demonstrated safety and effectiveness of AI-powered medical communication tools have important implications for healthcare policy and practice. Regulatory frameworks for AI health applications should consider communication tools as distinct from diagnostic or treatment AI, with appropriate oversight mechanisms that balance innovation with patient safety.

Healthcare organizations may benefit from pilot implementations of validated AI communication tools, particularly in settings serving diverse patient populations or those with known health literacy challenges. Integration with existing patient portal systems could provide seamless access while maintaining clinical oversight.

Professional medical societies should consider guidelines for AI communication tool use, including recommendations for clinical review processes and patient education about AI-generated content.

## Conclusions

As healthcare increasingly embraces digital transformation, validated AI communication tools represent a promising avenue for improving patient understanding, engagement, and health outcomes. This comprehensive validation study demonstrates that AI can safely and effectively personalize medical communication while maintaining clinical accuracy and safety. The Patiently AI application achieved substantial readability improvements, high expert ratings, and strong patient acceptance across diverse medical scenarios and target audiences. These findings support the potential for AI-powered tools to address persistent health literacy challenges at scale. However, successful implementation will require appropriate clinical oversight, continued safety monitoring, and integration with existing healthcare workflows.

## Data Availability

All data produced in the present study are available at the Open Science Framework repository: https://doi.org/10.17605/OSF.IO/3CDX6

## Acknowledgements

I thank the healthcare professionals who provided expert review and the patients who participated in evaluation surveys. No external funding was used for this study.

## Author contributions

N.L. conceived the study, developed the mobile application, designed the experiments, collected and analyzed the data, and wrote the manuscript.

## Data availability

Anonymized datasets supporting this study are available in the Open Science Framework repository at https://doi.org/10.17605/OSF.IO/3CDX6.

## Additional information

## References

1. Institute of Medicine. Health Literacy: A Prescription to End Confusion. Washington, DC: The National Academies Press; 2004.

2. NHS England. Improving health literacy. Available at: https://www.hee.nhs.uk/our-work/knowledge-library-services/improving-health-literacy. Accessed 14 September 2025.

3. Berkman, N. D. et al. Low health literacy and health outcomes: an updated systematic review. Ann. Intern. Med. 155, 97–107 (2011)

4. Baker, D. W. et al. Functional health literacy and the risk of hospital admission among Medicare managed care enrollees. Am. J. Public Health. 92, 1278–1283 (2002).

5. Schillinger, D. et al. Association of health literacy with diabetes outcomes. JAMA. 288, 475– 482 (2002).

6. Vernon, J. A., Trujillo, A., Rosenbaum, S. & DeBuono, B. Low health literacy: Implications for national health policy. Health Policy and Management Faculty Publications (2007).

7. 21st Century Cures Act, H.R. 34, 114th Congress (2016).

8. Pyper, C., Amery, J., Watson, M., Crook, C. Patients’ experiences when accessing their online electronic patient records in primary care. Br. J. Gen. Pract. 54, 38–43 (2004).

9. Schwartzberg, J. G., Cowett, A., VanGeest, J., Wolf, M. S. Communication techniques for patients with low health literacy: a survey of physicians, nurses, and pharmacists. Am. J. Health Behav. 31, S96–S104 (2007).

10. Wilson-Stronks A et al. One Size Does Not Fit All: Meeting the Health Care Needs of Diverse Populations. Oakbrook Terrace, IL: The Joint Commission; 2008.

11. Lee, P., Bubeck, S., Petro, J. Benefits, limits, and risks of GPT-4 as an AI chatbot for medicine. N. Engl. J. Med. 388, 1233–1239 (2023).

12. Singhal, K. et al. Large language models encode clinical knowledge. Nature. 620, 172–180 (2023).

13. Zack, T. et al. Coding Inequity: Assessing GPT-4’s Potential for Perpetuating Racial and Gender Biases in Healthcare. medRxiv. 2023.

14. Johnson, D., et al. Assessing the accuracy and reliability of AI-generated medical responses: an evaluation of the Chat-GPT model. Res Sq. 2023.

15. Patel, S. B., Lam, K. ChatGPT: the future of discharge summaries? Lancet Digit. Health. 5, e107–e108 (2023).

16. Ayers, J. W. et al. Comparing physician and artificial intelligence chatbot responses to patient questions posted to a public social media forum. JAMA Intern. Med. 183, 589–596 (2023).

17. Chen, S. et al. Use of artificial intelligence chatbots for cancer treatment information. JAMA Oncol. 9, 1459–1462 (2023).

18. Stanceski, K. et al. The quality and safety of using generative AI to produce patient-centred discharge instructions. npj Digit. Med. 7, 329 (2024).

19. Rudd, R. E., Anderson, J. E. The Health Literacy Environment of Hospitals and Health Centers. Boston, MA: Harvard School of Public Health; 2006.

20. Nielsen-Bohlman, L., Panzer, A. M, Kindig, D. A, editors. Health Literacy: A Prescription to End Confusion. Washington, DC: National Academies Press; 2004.

21. Abrams, M. A., Klass, P., Dreyer, B. P. Health literacy and children: introduction. Pediatrics. 124, S262–S264 (2009).

22. Manganello, J. A. Health literacy and adolescents: a framework and agenda for future research. Health Educ. Res. 23, 840–847 (2008).

23. Street, R. L., Makoul, G., Arora, N. K. & Epstein, R. M. How does communication heal? Pathways linking clinician–patient communication to health outcomes. Patient Educ. Couns. 74, 295–301 (2009).

24. Liu, X. et al. Reporting guidelines for clinical trial reports for interventions involving artificial intelligence: the CONSORT-AI extension. Lancet Digit. Health. 2, e537–e548 (2020).

25. Doak, C. C., Doak, L. G., Root, J.H. Teaching Patients with Low Literacy Skills. 2nd ed. Philadelphia, PA: JB Lippincott; 1996.

26. Shah, SJ et al. Clinician Perspectives on AI-Generated Drafts of Patient Test Result Explanations. JAMA Network Open. 8, e2528794 (2025).

27. Stellefson, M et al. eHealth literacy among college students: a systematic review with implications for eHealth education. J Med. Internet Res. 13, e102 (2011).

28. Pal, A et al. Generative AI/LLMs for Plain Language Medical Information for Patients, Caregivers and General Public: Opportunities, Risks and Ethics. Patient Prefer. Adherence. 31, 2227–2249 (2025).

